# Effect of oral contraceptive consumption timing on substrate metabolism, cognition, and exercise performance in females: a randomised controlled trial

**DOI:** 10.1101/2024.09.17.24311270

**Authors:** D Martin, M Bargh, K Pennington

## Abstract

The pharmacokinetic profile of oral contraceptives (OCs) results in an acute, transient increase in circulating synthetic reproductive hormones. The aim of this study was to examine the effects of acute OC ingestion on cognitive function, substrate metabolism and exercise performance. Sixteen combined OC (30µg ethinyl oestradiol and 150mg Levonorgestrel) users ingested either their OC or placebo (PLA) in a randomised, double-blind, crossover manner. After 60 minutes, participants completed tests of verbal memory and verbal fluency, followed by sub-maximal treadmill exercise for 6 minutes at 60% lactate threshold (LT) and 90% LT where respiratory exchange ratio (RER), carbohydrate oxidation, fat oxidation, heart rate (HR), rating of perceived exertion (RPE), felt arousal and feeling scale were recorded. Participants then completed an incremental ramp test to exhaustion to assess time to exhaustion (TTE) and peak oxygen uptake (VO_2_peak), before ingesting the pill corresponding to the condition, they were not in. RER, arousal and feeling scale were all significantly lower in the OC condition compared to PLA (all P < 0.05) with no difference in HR, carbohydrate oxidation, fat oxidation, RPE, TTE or VO_2_peak (all P > 0.05). Verbal fluency score was significantly higher in the OC condition compared to PLA (P < 0.05), with no differences between conditions for any aspect of verbal memory (all P < 0.05). Combined OC ingestion acutely affects substrate metabolism, affective responses to exercise and verbal fluency. The timing of OC ingestion should be considered in relation to aspects of physiological function.

**Key findings:**

- Ingestion of a combined oral contraceptive (OC) results in a transient increase in circulating synthetic reproductive hormones.
- The altered reproductive hormone profile affects substrate metabolism and affective responses to exercise.
- Cognitive performance in a verbal fluency task is also improved post OC ingestion.
- The within-day timing of OC ingestion can be manipulated to affect aspects of physiology, which may have implications for athletes, exercises and the general population, in addition to interpretation of prior research using OCs.

## Introduction

Reproductive hormones have been shown to influence substrate utilisation at rest and during exercise (Devries et al., 2006), exercise performance (McNulty et al., 2020) and cognitive function (Luine, 2014). Given that most women of reproductive age have used at least one form of hormonal contraception (87.5%; Daniels et al., 2013) and athletic populations have a high prevalence (∼50%) of hormonal contraceptive use (Martin et al., 2018), it is important to understand the effects of exogenous/synthetic hormones contained within hormonal contraceptives on outcomes of importance for female athletes, exercisers, and the general population.

The effect of female reproductive hormones on substrate metabolism is evidenced by a greater fat oxidation in females compared to males (Devries et al., 2006; Zehnder et al., 2005), and fat oxidation fluctuating across the menstrual cycle with the highest values observed during the luteal phase (Hackney et al., 1994; Zderic et al., 2001). The administration of exogenous oestrogen to amenorrheic women has been shown to reduce carbohydrate oxidation (Ruby et al., 1997). Several studies have shown reduced carbohydrate oxidation, reduced blood glucose concentrations and increased growth hormone concentrations in combined OC users vs. non-users (Bemben et al., 1992; Bonen et al., 1991). Furthermore, Ortega-Santos et al. (2018) showed that fat oxidation during exercise was significantly higher during pill consumption compared to the pill-free interval, suggesting that the exogenous hormones contained within OCs may be beneficial for enhancing fat oxidation. However, not all findings support this (*e.g.,* Issaco et al. 2012) and research on the effects of the menstrual cycle and OC use on exercise performance is equivocal likely due to poor study quality and heterogenous research designs (Elliott-Sale et al., 2020; McNulty et al., 2020).

Oestrogen concentrations have also been positively associated with aspects of cognition such as verbal memory and verbal fluency (Luine, 2014), which are superior in women compared to men (Hirnstein et al., 2023; Voyer et al., 2021) and superior in phases of the menstrual cycle with elevated oestrogen concentrations (Luine, 2014; Maki et al., 2002; Rosenberg & Park, 2002; Solís-Ortiz & Corsi-Cabrera, 2008). A systematic review (Warren et al., 2014) identified that verbal memory is the cognitive domain most consistently improved with OC use. Verbal memory performance was superior in combined OC users compared to non-users (e.g., Gogos, 2013) and verbal memory and verbal fluency performance were superior on pill consumption days compared to the pill-free interval (Mordecai et al., 2008).

To date, research has primarily compared hormonal contraceptive users to non-users or compared across phases of the menstrual cycle or OC cycle to evaluate the effects of reproductive hormones, however this does not consider the pharmacokinetic profile and pharmacodynamics of OCs. Combined OCs, the most prevalent type of OC used in athletic populations (Martin et al., 2018), display typical first order pharmacokinetics, whereby peak blood levels of ethinyl oestradiol and progestins are observed 60-120 minutes following ingestion of OCs (Dibbelt et al., 1991; Westhoff et al., 2010) followed by an exponential elimination proportionate to the concentration, with near total elimination at trough concentration (24 h). As such, there is only a window of a few hours whereby concentrations of ethinyl oestradiol and progestins are significantly elevated post-ingestion. These synthetic hormones have been shown to have a greater affinity for oestrogen receptors (ER-α and ER-β) than naturally occurring oestrogens (Dickson & Eisenfeld, 1981; Jeyakumar et al., 2011) and have potent physiological effects such as increasing the risk of venous thromboembolism (van Hylckama Vlieg et al., 2009) and breast cancer (Beaber et al., 2014), as well as reducing the risk of developing ovarian cancer (Iversen et al., 2018).

No research to date has explored the effect of this predictable, consistent and acute rise in exogenous hormones on aspects of physiology and cognitive function, potentially due to the ethical constraints of supplementing individual doses of OCs to non-users. The current study utilises a novel study design to circumvent these ethical constraints; by using a double-blind, placebo-controlled experiment whereby the timing of OC ingestion is manipulated in existing OC users to compare participants when they have peak and trough exogenous hormone concentrations circulating in their system. Utilising this novel design, the aim of this study was to assess whether the acute ingestion of a combined OC affected cognition, substrate metabolism and exercise performance, compared to the ingestion of a placebo. The primary hypotheses were that following OC ingestion, RER will be lower, and verbal memory and verbal fluency will be superior, compared to the placebo condition.

## Methods

### Participants

Twenty physically active female participants from the United Kingdom volunteered to take part in the study between January 2020 and December 2023. Following the withdrawal of 4 participants due to scheduling issues, 16 participants (age = 20.3 ± 2.3 y; height = 1.67 ± 0.08 m; body mass = 64.9 ± 11.2 kg) completed the study. An *a priori* sample size calculation using data from Hackney et al. (1994) identified that 12 participants are required for a power of β = 0.95, with respiratory exchange ratio (RER) as the main outcome measure and a sample size of 16 participants was needed for a power of β = 0.95 for verbal memory (Mordecai et al., 2008) therefore the trial was stopped upon completion of 16 participants. Participants were required to have used a combined oral contraceptive with 30 µg ethinyl oestradiol and 150 mg Levonorgestrel (Rigevidon®, n = 12; Microgynon®, n = 2; Maexeni® 30, n = 1; Levest®, n = 1) for at least 6 months prior to participation, with a regimen of 21 pill consumption days and a 7-day pill free interval. Further inclusion criteria were being aged 18-35 years, physically active, and speak fluent English. Exclusion criteria included polycystic ovarian syndrome, endometriosis, pregnancy, childbirth or lactation in the previous 6 months, BMI < 18.5 or > 30 kg·m^2^, smokers, any disorder known to affect metabolic health, history of head injury/neurological disorders and history of psychiatric disorders such as major depression or anxiety disorder. Ethical approval was received from the University of Lincoln Research Ethics Committee (718) and conformed to the standards of the Declaration of Helsinki. Participants were provided with a participant information sheet, completed a health screen, and gave their written informed consent in writing prior to commencing the study. Participants could withdraw from the study at any time.

### Experimental design

Participants completed a familiarisation session, followed by two main experimental trials in counterbalanced order in accordance with CONSORT guidelines (Schulz et al., 2010). Prior to the main trials, participants provided the technical team with two OCs from their pill packet to be allocated in a double-blind manner. The technical team conducted a simple randomisation (www.randomizer.org), ensuring allocation was independent of the research team. The allocation details were only revealed after the completion of data collection. Oral contraceptives and lactose placebos matched for colour, size and appearance were concealed in opaque cannisters with labels identifying allocation. In the OC main trial (OC), participants ingested their OC at 08:00 h, followed by the placebo at the completion of testing at approximately ∼10:00 h. In the placebo trial (PLA), participants ingested the placebo at 08:00 h, followed by their OC after testing had completed. Main experimental trials took place on OC consumption days within the same OC cycle, separated by no more than 10 days to minimise the effect of synthetic hormone accretion over a pill consumption phase (Kuhnz et al., 1992). Testing did not take place during the first 3 days of pill consumption due to the elevated endogenous oestrogen concentrations at this time (van Heusden & Fauser, 1999). Participants were asked to consume their OC at 08:00 on OC consumption days for at least 1 week before attending the laboratory for the main trials so that upon arrival to the laboratory they had not ingested their OC for 24 h. Participants were asked to avoid exercise and alcohol for 24 hours before the main trials and to attend the laboratory following an overnight fast with *ad libitum* water intake.

### Experimental protocol

In the familiarisation session, participants completed a practice trial of the Rey Auditory Verbal Learning Test (RAVLT) and Verbal Fluency task using alternative task forms to reduce practice effects (Bell et al., 2018). Participants then completed a two-stage lactate threshold and VO_2_peak test (Jones, 1998) to establish steady state exercise intensities for the sub-maximal exercise assessment in the main trial and to familiarise participants with the maximal exercise test.

In the main trial, participants attended the laboratory at 08:00 and either ingested their OC or PLA in a double-blind manner according to the randomly generated sequence. Participants were asked to open the concealed container and ingest the tablet with water and with their eyes closed to reduce any possibility of them distinguishing between the OC and PLA condition. Participants then rested for 60 minutes to allow time for the OC or PLA to enter the circulation, before completing the cognitive test battery.

### Cognitive tests

The cognitive test battery was performed in a quiet area, with participants seated at a desk facing a blank wall to minimise distractions. During the tests, the experimenter was seated approximately 1 m behind the participant and participants were instructed to face away from the experimenter throughout the tests.

Verbal memory was assessed using the RAVLT (Rey, 1941) to assess immediate memory span, new learning, and susceptibility to interference. A list of 15 words (List A) was read aloud for 5 consecutive trials (Trials 1-5), each followed by a free recall test during which participants were asked to recall as many words as possible from the word list. Following this, an interference list of 15 different words (List B) was read aloud, followed by a free recall of this list. Immediately after this, participants were asked to recall List A without further presentation of these words (Trial 6). After a 30-minute delay, the participants were then asked to recall List A without hearing the words again (Trial 7). Time limits were not imposed, and participants were asked to inform the experimenter when they could not remember any more words. Acquisition (sum of all words), learning rate (difference between Trial 1 and 5), proactive interference (difference between words recalled in Trial 1 and List B), retroactive interference (difference between Trial 5 and 6) and forgetting (difference between Trial 7 and 5) were calculated. Alternate word lists with equivalent word length, serial position and frequency in the English language (Geffen et al. 1994) were counter-balanced across main trials.

The Verbal fluency test (Benton, 1989) is an assessment of the ability to produce words given certain restrictions. In this test, participants were given a letter of the alphabet and were asked to say as many words as possible that begin with that letter in one minute, excluding capitalised words (*i.e.,* proper nouns) or variations of the same word (*e.g.,* garden and gardening). In each assessment, participants were given three different letters, with a short break between each. The total number of acceptable words said within the time limit was recorded with higher numbers equating to greater verbal fluency. The letter combinations used have been shown to have high equivalence (Cohen & Stanczak, 2000) and the order of letter combinations was counter-balanced across main trials.

In addition to the above tests, a laptop-based cognitive testing battery was completed which included the mental rotation test (Vandenberg & Kuse, 1978), Stroop-colour test (Stroop, 1935), Corsi blocks test (Corsi, 1972) and Rapid Visual Information Processing (RVIP) task (Wesnes & Warburton, 1984). However, a technical issue resulted in data loss for 7 participants in these tests and therefore data are only available for 9 participants. We have not included these data in the paper as they are not sufficiently powered.

### Exercise tests

Immediately after the cognitive tests (∼100 minutes post OC/PLA ingestion), participants completed a sub-maximal exercise assessment on a treadmill (Mercury, h/p/cosmos, Germany) for 6 minutes at 60%LT and 6 minutes at 90%LT. Exercise intensities below LT were used as previous research has shown that during exercise at an intensity greater than LT, the demand for increased carbohydrate consumption surpasses the influence of oestrogen (Hackney, 2021; Hackney et al., 1994). In the last minute of each stage, metabolic variables were collected using breath-by-breath analysis (Metalyser 3B, Cortex, Germany) and heart rate, breathing frequency, rating of perceived exertion (Borg, 1970), felt arousal (Svebak & Murgatroyd, 1985) and feeling scale (Hardy & Rejeski, 1989) were recorded. Respiratory exchange ratio (RER), carbohydrate oxidation and fat oxidation (Frayn, 1983) and energy expenditure were calculated from the metabolic data. Immediately following the sub-maximal exercise test, participants completed a maximal treadmill ramp test at the same starting speed as the familiarisation, increasing the incline by 1% each minute until volitional exhaustion. Time to exhaustion, maximal heart rate and VO_2_peak were recorded. Following the completion of the exercise test, participants ingested the tablet (OC or PLA) that they had not ingested upon arrival, maintaining the double-blind design. No harms or unintended effects were reported for any participant.

### Statistical analysis

Heart rate, breathing frequency, RER, carbohydrate oxidation, fat oxidation, energy expenditure, RPE, arousal and feeling scale were compared between conditions (OC vs. PLA) and exercise intensities (60% vs. 90% LT) using two-way repeated measures ANOVAs, with Bonferroni correction. RAVLT outcomes, verbal fluency score, VO_2_peak, time to exhaustion and maximum HR during the ramp test were compared using paired-samples t-tests after checking for normality using Shapiro-Wilk tests. Data are presented as mean ± SD and statistical significance was set at P ≤ 0.05.

## Results

### Sub-maximal exercise

For RER, there was a significant effect of intensity (F(1, 15) = [151.68], p < 0.001, ηp^2^ = 0.910) and condition (F(1,15) = [7.90], p = 0.0132, ηp^2^ = 0.345) with no intensity x condition interaction (F(1,15) = [1.35], p = 0.263, ηp^2^ = 0.083). At 60%LT, RER was significantly lower in OC (0.855 ± 0.044) compared to PLA (0.871 ± 0.041, p = 0.0376, D = 0.37). AT 90%LT, RER was significantly lower in OC (0.917 ± 0.041) compared to PLA (0.939 ± 0.034, p = 0.0369, D = 0.59; Figure 1). For heart rate, breathing frequency, carbohydrate oxidation, fat oxidation and energy expenditure there was a significant effect of intensity (all p < 0.05), with no condition or intensity x condition interaction (all > 0.05; Table 1).

**Figure 1.**
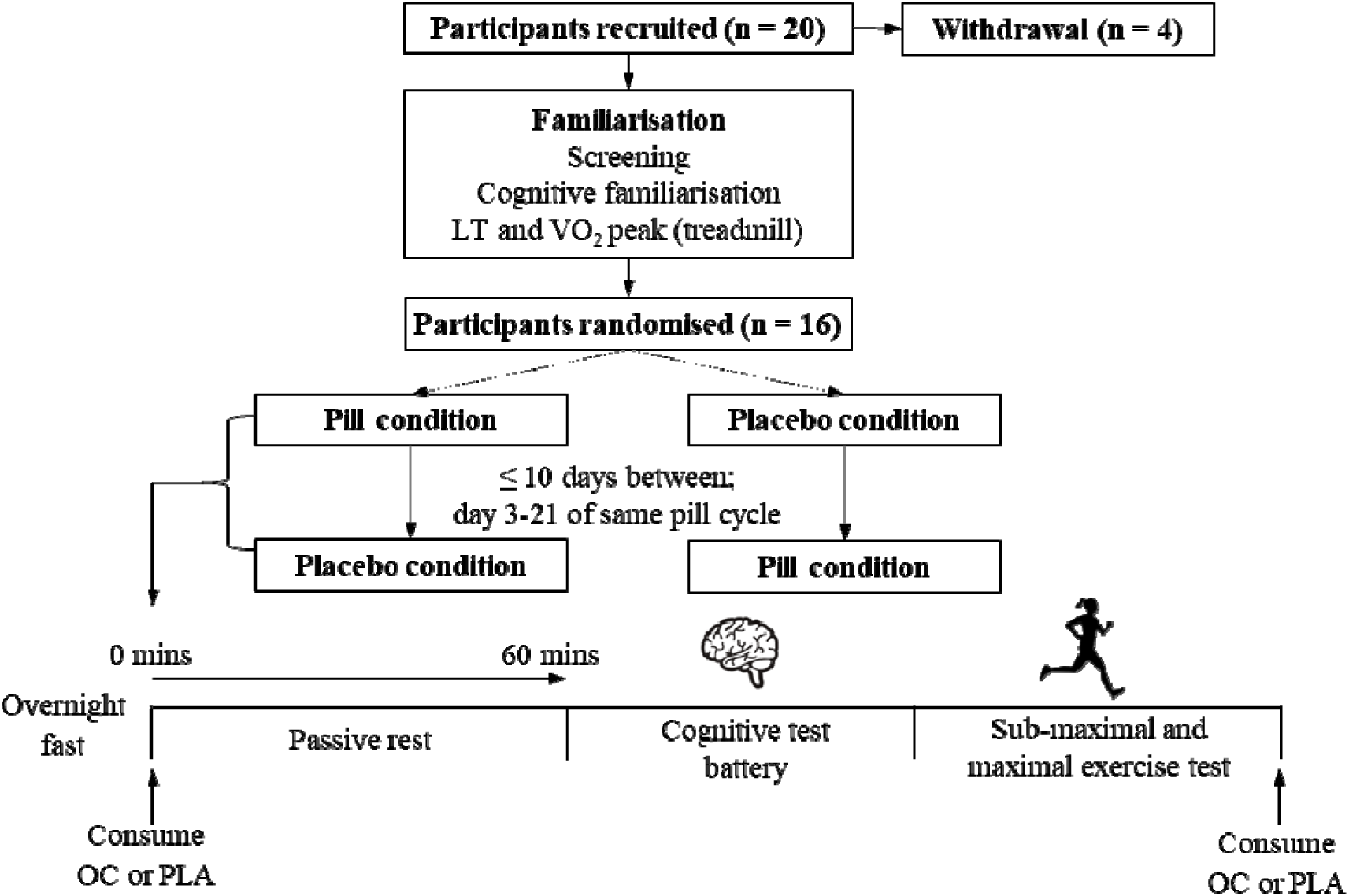
Schematic of the study. Lactate threshold; LT, Peak oxygen uptake; VO_2_peak; Oral contraceptive; OC, Placebo; PLA.

**Figure 1.**
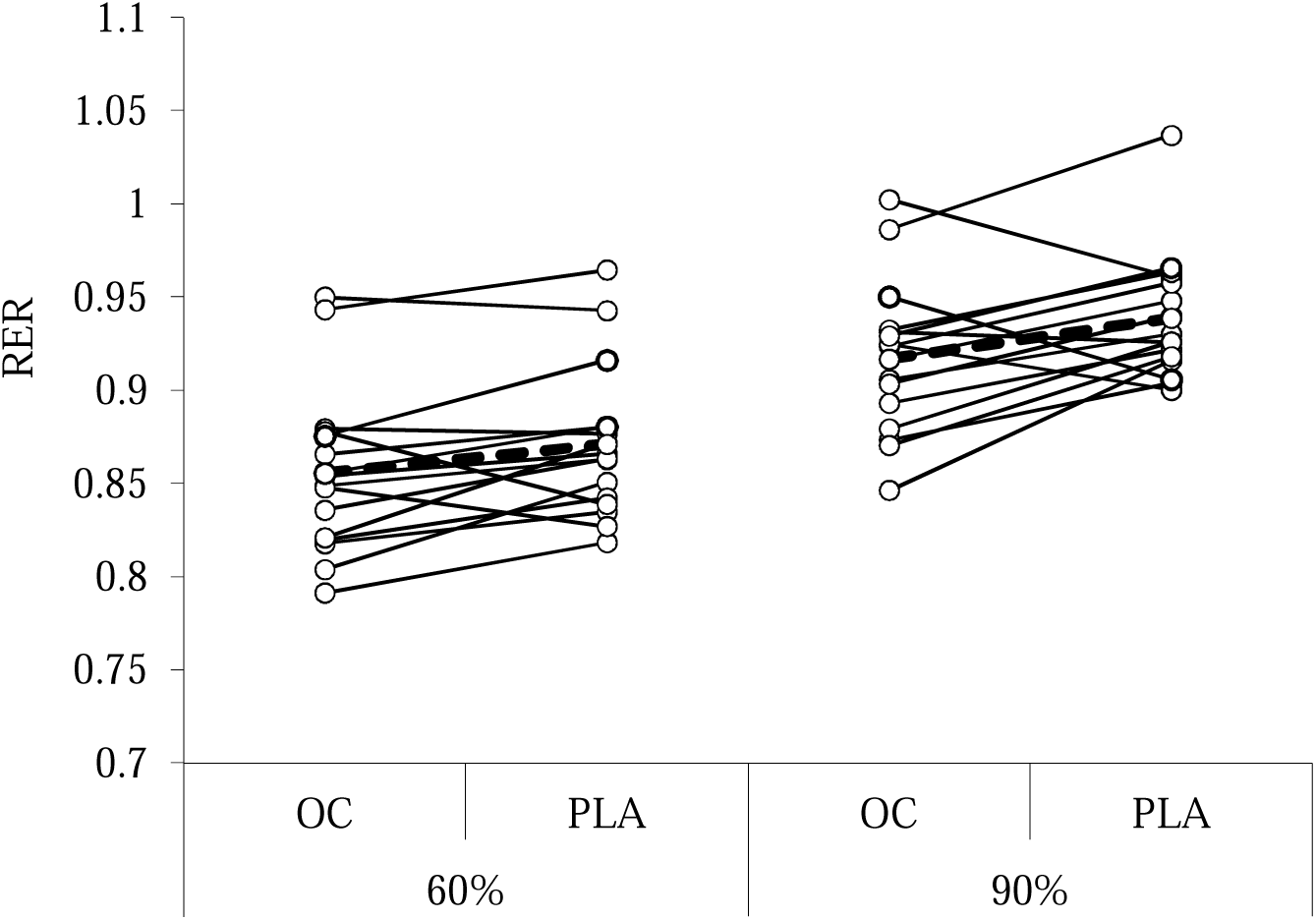
Univariate scatter plot with individual and mean Respiratory Exchange Ratio (RER) for Oral Contraceptive (OC) and Placebo (PLA) conditions at 60% and 90% lactate threshold. For arousal, there was a significant effect of condition (F(1,15), = [5.29], p = 0.0362, ηp^2^ = 0.261), with no significant effect of intensity (F(1, 15), = [1.52], p = 0.237, ηp^2^ = 0.092) or intensity x condition interaction (F(1,15), = [1.90], p = 0.188, ηp^2^ = 0.113). Post-hoc tests showed arousal was significantly lower (p = 0.0280, D = 0.39) in OC (2.13 ± 1.15) compared to PLA (2.56 ± 1.09) at 60%LT, with no significant difference at 90%LT (p > 0.05). For feeling scale, there was a significant effect of condition (F(1,15), = [8.23], p = 0.0117, ηp^2^ = 0.354), with no significant effect of intensity (F(1, 15), = [3.92], p = 0.0664, ηp^2^ = 0.207) or intensity x condition interaction (F(1,15), = [3.85], p = 0.07, ηp^2^ = 0.204). Post-hoc tests showed feeling was significantly (p = 0.026, D = 0.47) lower in OC (1.75 ± 1.61) compared to PLA (2.50 ± 1.55) at 60%LT, with no significant difference at 90%LT (P > 0.05). For RPE, there was a significant effect of intensity (F(1, 15), = [34.92], p < 0.001, ηp^2^ = 0.700), with no condition (F(1,15), = [0.03], p = 0.872, ηp^2^ = 0.002) or intensity x condition interaction (F(1,15), = [0.18], p = 0.676, ηp^2^ = 0.012; Table 1).

**Table 1.** Mean ± SD measures during sub-maximal exercise for Oral Contraceptive (OC) and Placebo (PLA) conditions at 60% and 90% lactate threshold.

|  | 60% |  | 90% |  |
| --- | --- | --- | --- | --- |
|  | OC | PLA | OC | PLA |
| $\text{VO}_2$ ($\text{L}\cdot\text{min}^{-1}$) | $0.93 \pm 0.17$ | $0.96 \pm 0.23$ | $1.28 \pm 0.31$ | $1.29 \pm 0.31$ |
| $\text{VCO}_2$ ($\text{L}\cdot\text{min}^{-1}$) | $0.80 \pm 0.17$ | $0.83 \pm 0.20$ | $1.17 \pm 0.31$ | $1.21 \pm 0.30$ |
| RER | $0.86 \pm 0.04$ | $0.87 \pm 0.04$ | $0.92 \pm 0.04$ | $0.94 \pm 0.03$ |
| Carbohydrate oxidation ($\text{g}\cdot\text{min}^{-1}$) | $0.64 \pm 0.28$ | $0.70 \pm 0.24$ | $1.24 \pm 0.45$ | $1.35 \pm 0.41$ |
| Fat oxidation ($\text{g}\cdot\text{min}^{-1}$) | $0.22 \pm 0.07$ | $0.21 \pm 0.09$ | $0.17 \pm 0.09$ | $0.14 \pm 0.11$ |
| Energy expenditure ($\text{kJ}\cdot\text{min}^{-1}$) | $27.6 \pm 34.9$ | $19.7 \pm 4.7$ | $26.4 \pm 6.5$ | $27.00 \pm 6.58$ |
| Breathing frequency ($\text{breaths}\cdot\text{min}^{-1}$) | $26.6 \pm 6.2$ | $27.1 \pm 6.4$ | $33.9 \pm 7.7$ | $35.5 \pm 7.5$ |
| Heart rate ($\text{beats}\cdot\text{min}^{-1}$) | $113.3 \pm 12.6$ | $114.3 \pm 13.4$ | $142.4 \pm 20.1$ | $143.2 \pm 19.6$ |
| RPE | $8.4 \pm 1.8$ | $8.3 \pm 1.7$ | $10.1 \pm 2.3$ | $10.3 \pm 2.0$ |
| Arousal | $2.1 \pm 1.1$ | $2.6 \pm 1.1$ | $2.4 \pm 1.1$ | $2.6 \pm 1.2$ |
| Feeling scale | $1.8 \pm 1.6$ | $2.5 \pm 1.5$ | $1.6 \pm 1.5$ | $1.9 \pm 1.6$ |

### Ramp test measures

There was no significant difference between OC and PLA for VO_2_peak (P = 0.0987; D = 0.44) time to exhaustion (P = 0.461, D = 0.189) or HR_max_ (P = 0.172, D = 0.357; Table 2).

**Table 2.** Mean ± SD ramp test measures and cognitive performance

| Measure | OC | PLA |
| --- | --- | --- |
| *Ramp test outcomes* |  |  |
| VO<sub>2</sub>peak (L·min<sup>-1</sup>) | 2.18 ± 0.334 | 2.13 ± 0.360 |
| TTE (s) | 468.8 ± 78.1 | 463.9 ± 83.6 |
| HR<sub>max</sub> (beats·min<sup>-1</sup>) | 196.2 ± 7.0 | 195.0 ± 7.4 |
| *Cognitive performance* |  |  |
| Acquisition | 84.6 ± 15.0 | 85.9 ± 16.1 |
| Learning rate | 5.1 ± 2.1 | 5.3 ± 2.5 |
| Proactive interference | 1.3 ± 1.7 | 1.3 ± 2.5 |
| Retroactive interference | 1.7 ± 2.8 | 1.8 ± 1.4 |
| Forgetting | 2.1 ± 2.8 | 2.9 ± 2.3 |
| Verbal fluency | 45.9 ± 12.7 | 40.7 ± 8.3* |

\*Indicates significant difference between OC and PLA,

### Verbal cognitive tests

There were no significant differences between OC and PLA conditions (all P > 0.5) for RAVLT acquisition, learning rate, proactive interference, retroactive interference and forgetting. Verbal fluency performance was significantly greater (P = 0.047; D = 0.54) in OC (45.9 ± 12. 7) compared to PLA (40.7 ± 8.3); Table 2).

## Discussion

The aim of this study was to assess whether the acute ingestion of a combined OC affected cognition, substrate metabolism and exercise performance, compared to the ingestion of a placebo. In agreement with the primary hypotheses, RER was lower and verbal fluency was superior following OC ingestion compared to PLA, however there was no difference in verbal memory performance between the conditions. There was also no difference in exercise performance or VO_2_ peak between conditions, however felt arousal and feeling scale were lower in the OC condition.

The lower RER observed during sub-maximal exercise following OC ingestion compared to a placebo is similar to that observed in studies of the menstrual cycle, whereby RER was lower in the luteal phase compared to the follicular phase (Hackney et al., 1994; Wenz et al., 1997; Zderic et al., 2001). A lower RER is indicative of a greater proportional use of fat as a fuel, which is in-line with the proposed mechanisms of oestrogen on substrate metabolism. Oestrogen influences substrate metabolism indirectly via actions on growth hormone and insulin (D’Eon & Braun, 2002) and directly via glucose transporter-4 (GLUT4) translocation (Campello et al., 2017; Yue et al., 2020) and increased lipoprotein lipase and hormone-sensitive lipase activity (Campbell & Febbraio, 2001; D’Eon et al., 2005) among other mechanisms (for review see Oosthuyse et al., 2023). It has been hypothesised that the oral delivery of combined OCs results in high hepatic exposure via the portal vein which could exacerbate the effects of these hormones in hepatocytes (Magkos et al., 2022) thereby resulting in marked changes to substrate metabolism. Increased oestrogen receptor mediated 5’AMP-activated (AMPK) phosphorylation activity occurs within 10 minutes of oestrogen administration (D’Eon et al., 2008; Gorres et al., 2011) demonstrating that the changes in metabolism observed with the transient spike in exogenous hormones in the current study are plausible.

In contrast to some studies of sex or menstrual cycle effects on substrate metabolism during exercise (e.g., Wenz et al. 1997), the current study did not find significant differences between conditions for either fat or carbohydrate oxidation rate, despite significant differences in RER.. However, as RER is a ratio of two correlated values (VO_2,_ VCO_2_), the pooled coefficient of variation was much smaller (4.9%) compared to carbohydrate (38.09%) and fat (36.3%) oxidation rates, thereby improving the likelihood of statistical significance, a known issue with ratio data (Jasienski & Bazzaz, 1999). The absence of significant differences in fat and carbohydrate oxidation between the OC and placebo condition may also be related to type and timing of reproductive hormone concentrations used in the current study. Prior research has shown that the change in fat oxidation from the early follicular phase to mid-luteal phase was positively correlated with the change in oestrogen to progesterone ratio (Hackney et al., 2022), potentially as a result of the anti-oestrogenic effect of progesterone (Hatta et al., 1988). Specifically, progesterone has been shown to repress oestrogen-mediated GLUT4 translocation (Campbell & Febbraio, 2002). Participants in the current study all used the same type and concentration of progestin (150 mg Levonorgestrel) which might elicit different progestogenic effects compared to endogenous progesterone or other types (*i.e.,* generations) and concentrations of progestins in other hormone therapies (Mitchell & Welling, 2020). Furthermore, 30µg ethinyl oestradiol was used by all participants which might have different physiological effects compared to other types of oestrogen (e.g., oestradiol or oestradiol valerate) or doses of ethinyl oestradiol used in many hormonal contraceptives (20-50 µg; Stanczyk et al., 2013). Further work is needed to explore the pharmacodynamic effect of different concentrations and types of hormonal contraceptives (e.g. progestin-only contraceptives), in addition to other hormonal therapies such as hormone replacement therapy (HRT).

Concomitant with a greater proportional use of fat as a fuel in the OC condition, was a lower felt arousal and feeling scale score, which are considered together as measures of affective valence (Bastos et al., 2023). Reduced affective valence in the presence of higher oestrogen concentrations has previously been observed when exercising in the luteal phase of the menstrual cycle (Prado et al., 2021) although the mechanisms behind this are unclear. Glucose availability is positively associated with affective valence (Backhouse et al., 2007; Lee et al., 2018) so it may be that lower proportional carbohydrate oxidation resulted in lower affective valence. Alternatively, OC use blunts the cortisol response to exercise (Kirschbaum et al., 1996) and down-regulates IGF-1 concentrations which mediate the positive effects of exercise on mood in females (Munive et al., 2016). Oestrogen may also directly attenuate arousal via cortical-sub-cortical control within hypothalamic-pituitary-adrenal circuitry (Goldstein et al., 2005). Typically, lower affective valences are observed with greater exercise intensity (Kilpatrick et al., 2007) or perceived exertion (Farias-Junior et al., 2020), however treadmill speed was identical between conditions and no significant differences in RPE were observed. Experiencing positive affective responses to exercise makes individuals more likely to continue to undertake exercise (Rhodes & Kates, 2015) and therefore the timing of OC consumption around exercise should be researched in relation to enjoyment and participation.

Perceived exertion regulates the termination of exercise (Marcora & Staiano, 2010) and, in-line with the similar RPE scores during sub-maximal exercise, there were no significant differences between the OC and placebo conditions for time to exhaustion in the ramp test, VO_2peak_ or peak heart rate. It is purported that sex-hormone related differences are more likely to impact exercise performance in longer-duration exercise, whereby glycogen sparring via increased fat oxidation may confer a performance advantage (Tiller et al., 2021). Therefore, future research exploring the effects of OC consumption timing on exercise performance should include longer-duration exercise. Additionally, further research is required to consider the effects of OC consumption timing on other key metrics which may be mediated by sex-hormones such as injury risk (Barlow et al., 2024; Martin et al., 2018)

For the cognitive data, verbal fluency was superior in the OC condition compared to the placebo which is in-line with prior research showing improved performance in women compared to men, and in the phases of the menstrual cycle associated with the highest oestrogen concentrations (Luine, 2014). Griksiene and Ruksenas (2011) showed that participants using OCs containing third generation progestins (*e.g.,* gestodene, desogestrel and norgestimate) had poorer verbal fluency compared to newer, less androgenic OCs (e.g., drospirenone). The current study used 2^nd^ generation OCs (150mg levonorgestrel) which are similar in androgenicity to 3^rd^ generation OCs (Louw-du Toit et al., 2017) and therefore the pharmacodynamic effects observed in the current study may be different compared other OC formulations, especially considering levonorgestrel is one of few progestins which are oestrogen-receptor alpha (ERα) agonists (Low-du Toit et al. 2017). The current study found no effect of acute OC ingestion on verbal memory, despite this often being affected by alterations to reproductive hormone concentrations such as with the menstrual cycle or oral contraceptive use (Luine, 2014), although further research is required exploring other OC formulations and other exogenous hormone models.

### Limitations

The study protocol included the use of a computer-based cognitive test battery comprised of the mental rotation test, Stroop task, Corsi blocks test and RVIP test. However, due to a technical issue with the software, we only had complete data sets for 9 participants and therefore have removed these data from the study as these data were underpowered. Further research would benefit from studying the acute effects of OC ingestion on a wider range of cognitive tasks, especially the mental rotation test which is often affected by the reproductive hormone milieu (Peragine et al., 2020). We also collected blood samples for assessment of serum ethinyl oestradiol and levonorgestrel, however a storage issue meant that these data could not be used. Future research employing this design could collect serum samples to compare the pharmacokinetic profiles of the OCs between participants and examine whether this influences the pharmacodynamic response. Whilst an *a priori* power analysis was conducted for primary variables of interest (RER and verbal memory), we did not conduct power calculations for secondary variables and therefore some inferences in the current study may be underpowered. Lastly, we only included OC formulations with 30µg ethinyl oestradiol and 150mg levonorgestrel and therefore further research is required to explore other OC types (e.g., progesterone-only) formulations (i.e., different progestins) and concentrations, in addition to other exogenous reproductive hormone models such as hormone replacement therapy in menopausal women.

### Implications of research

This study has shown that the acute ingestion of OCs affects substrate metabolism, affective responses to exercise and verbal fluency and therefore the timing of OC ingestion may need to be considered in relation to these factors. Whilst additional research is required to corroborate these findings and identify other factors which may be affected by the timing of OC ingestion, this could impact how OCs are used in sport, exercise or professional settings where ergogenic effects are sought. It is important to highlight that OCs are primarily used to prevent pregnancy and that manipulating OC ingestion timings might increase the risk of ovulation and pregnancy potential. A systematic review identified that missing one to four consecutive OCs during the pill consumption phase resulted in little follicular activity and low risk of ovulation across 10 studies (Zapata et al., 2013). The current study manipulated the timing of OC ingestion by approximately 2 hours and guidance contained within OC leaflets indicates that you are protected from pregnancy if one OC is 12 hours late or less. Therefore, the proposed manipulation of OC timing is well within the bounds of acceptability and does not pose risk of reduced contraceptive efficacy.

Importantly, the current study has implications for existing research that has been conducted in OC users as very little research has standardised the timing of OC ingestion in relation to measurements. If studies are looking to compare OC users to non-users, or compare the phases of OC use, studies that do not consider the timing of ingestion of OC in relation to measurements may introduce a source of variability. The timing of OC ingestion should therefore be considered as a factor which could be controlled or considered in research as with other factors (e.g., Merrel et al., 2024), and evaluations of extant research in OCs users should be cautious of whether OC ingestion timing was controlled and the potential effects on research quality.

## Conclusion

The pharmacokinetic profile of OCs results in a transient increase in circulating concentrations of synthetic reproductive hormones, which can acutely affect substrate metabolism during exercise, affective responses to exercise, and verbal fluency performance. This is the first study to show that the timing of OC ingestion can affect aspects of physiological function.

## Declaration

This project was supported by internal funding from the University of Lincoln.

## Registration

Registered on Open Science Framework https://osf.io/m9jr3

## Author contributions

**DM:** Conceptualization, methodology, validation, formal analysis, investigation, resources, data curation, writing-original draft, visualization, project administration, funding acquisition. **MB:** Investigation, writing – review & editing. **KP:** Conceptualisation, methodology, writing – review & editing, funding acquisition

## Supporting information

CONSORT checklist

## Data Availability

All data produced in the present study are available upon reasonable request to the authors

## References

1. Backhouse, S. H., Ali, A., Biddle, S. J., & Williams, C. (2007). Carbohydrate ingestion during prolonged high-intensity intermittent exercise: impact on affect and perceived exertion. Scand J Med Sci Sports, 17(5), 605–610. 10.1111/j.1600-0838.2006.00613.x

2. Barlow, A., Blodgett, J. M., Williams, S., Pedlar, C. R., & Bruinvels, G. (2024). Injury Incidence, Severity, and Type Across the Menstrual Cycle in Female Footballers: A Prospective Three Season Cohort Study. Med Sci Sports Exerc, 56(6), 1151–1158. 10.1249/mss.0000000000003391

3. Bastos, V., Rodrigues, F., Davis, P., & Teixeira, D. S. (2023). Assessing affective valence and activation in resistance training with the feeling scale and the felt arousal scale: A systematic review. PLoS One, 18(11), e0294529. 10.1371/journal.pone.0294529

4. Beaber, E. F., Buist, D. S., Barlow, W. E., Malone, K. E., Reed, S. D., & Li, C. I. (2014). Recent oral contraceptive use by formulation and breast cancer risk among women 20 to 49 years of age. Cancer Res, 74(15), 4078–4089. 10.1158/0008-5472.Can-13-3400

5. Bell, L., Lamport, D. J., Field, D. T., Butler, L. T., & Williams, C. M. (2018). Practice effects in nutrition intervention studies with repeated cognitive testing. Nutr Healthy Aging, 4(4), 309–322. 10.3233/nha-170038

6. Bemben, D. A., Boileau, R. A., Bahr, J. M., Nelson, R. A., & Misner, J. E. (1992). Effects of oral contraceptives on hormonal and metabolic responses during exercise. Med Sci Sports Exerc, 24(4), 434–441.

7. Benton, A., Hamsher, K., & Sivan, A. (1989). Multilingual Aphasia (3rd ed.). AJA Associates.

8. Bonen, A., Haynes, F. W., & Graham, T. E. (1991). Substrate and hormonal responses to exercise in women using oral contraceptives. J Appl Physiol (1985), *70*(5), 1917-1927. 10.1152/jappl.1991.70.5.1917

9. Borg, G. (1970). Perceived exertion as an indicator of somatic stress. Scand J Rehabil Med, 2(2), 92–98.

10. Campbell, S. E., & Febbraio, M. A. (2001). Effect of ovarian hormones on mitochondrial enzyme activity in the fat oxidation pathway of skeletal muscle. American Journal of Physiology-Endocrinology and Metabolism, 281(4), E803–E808. 10.1152/ajpendo.2001.281.4.E803

11. Campbell, S. E., & Febbraio, M. A. (2002). Effect of the ovarian hormones on GLUT4 expression and contraction-stimulated glucose uptake. Am J Physiol Endocrinol Metab, 282(5), E1139–1146. 10.1152/ajpendo.00184.2001

12. Campello, R. S., Fátima, L. A., Barreto-Andrade, J. N., Lucas, T. F., Mori, R. C., Porto, C. S., & Machado, U. F. (2017). Estradiol-induced regulation of GLUT4 in 3T3-L1 cells: involvement of ESR1 and AKT activation. J Mol Endocrinol, 59(3), 257–268. 10.1530/jme-17-0041

13. Cohen, M. J., & Stanczak, D. E. (2000). On the reliability, validity, and cognitive structure of the Thurstone Word Fluency Test. Arch Clin Neuropsychol, 15(3), 267–279.

14. Corsi. (1972). *Memory and the Medial Temporal Region of the Brain* McGill University].

15. D’Eon, T., & Braun, B. (2002). The roles of estrogen and progesterone in regulating carbohydrate and fat utilization at rest and during exercise. J Womens Health Gend Based Med, 11(3), 225–237. 10.1089/152460902753668439

16. D’Eon, T. M., Rogers, N. H., Stancheva, Z. S., & Greenberg, A. S. (2008). Estradiol and the estradiol metabolite, 2-hydroxyestradiol, activate AMP-activated protein kinase in C2C12 myotubes. Obesity (Silver Spring), 16(6), 1284–1288. 10.1038/oby.2008.50

17. D’Eon, T. M., Souza, S. C., Aronovitz, M., Obin, M. S., Fried, S. K., & Greenberg, A. S. (2005). Estrogen regulation of adiposity and fuel partitioning. Evidence of genomic and non-genomic regulation of lipogenic and oxidative pathways. J Biol Chem, 280(43), 35983–35991. 10.1074/jbc.M507339200

18. Devries, M. C., Hamadeh, M. J., Phillips, S. M., & Tarnopolsky, M. A. (2006). Menstrual cycle phase and sex influence muscle glycogen utilization and glucose turnover during moderate-intensity endurance exercise. Am J Physiol Regul Integr Comp Physiol, 291(4), R1120–1128. 10.1152/ajpregu.00700.2005

19. Dibbelt, L., Knuppen, R., Jütting, G., Heimann, S., Klipping, C. O., & Parikka-Olexik, H. (1991). Group comparison of serum ethinyl estradiol, SHBG and CBG levels in 83 women using two low-dose combination oral contraceptives for three months. Contraception, 43(1), 1–21. 10.1016/0010-7824(91)90122-v

20. Dickson, R. B., & Eisenfeld, A. J. (1981). 17 Alpha-ethinyl estradiol is more potent than estradiol in receptor interactions with isolated hepatic parenchymal cells. Endocrinology, 108(4), 1511–1518. 10.1210/endo-108-4-1511

21. Elliott-Sale, K. J., McNulty, K. L., Ansdell, P., Goodall, S., Hicks, K. M., Thomas, K., Swinton, P. A., & Dolan, E. (2020). The Effects of Oral Contraceptives on Exercise Performance in Women: A Systematic Review and Meta-analysis. Sports Med, 50(10), 1785–1812. 10.1007/s40279-020-01317-5

22. Farias-Junior, L. F., Browne, R. A. V., Astorino, T. A., & Costa, E. C. (2020). Physical activity level and perceived exertion predict in-task affective valence to low-volume high-intensity interval exercise in adult males. Physiology & Behavior, 224, 112960. 10.1016/j.physbeh.2020.112960

23. Frayn, K. N. (1983). Calculation of substrate oxidation rates in vivo from gaseous exchange. J Appl Physiol Respir Environ Exerc Physiol, 55(2), 628–634. 10.1152/jappl.1983.55.2.628

24. Gogos, A. (2013). Natural and synthetic sex hormones: Effects on higher-order cognitive function and prepulse inhibition. Biological Psychology, 93(1), 17–23. 10.1016/j.biopsycho.2013.02.001

25. Goldstein, J. M., Jerram, M., Poldrack, R., Ahern, T., Kennedy, D. N., Seidman, L. J., & Makris, N. (2005). Hormonal cycle modulates arousal circuitry in women using functional magnetic resonance imaging. J Neurosci, 25(40), 9309–9316. 10.1523/jneurosci.2239-05.2005

26. Gorres, B. K., Bomhoff, G. L., Morris, J. K., & Geiger, P. C. (2011). In vivo stimulation of oestrogen receptor α increases insulin-stimulated skeletal muscle glucose uptake. J Physiol, 589(Pt 8), 2041–2054. 10.1113/jphysiol.2010.199018

27. Griksiene, R., & Ruksenas, O. (2011). Effects of hormonal contraceptives on mental rotation and verbal fluency. Psychoneuroendocrinology, 36(8), 1239–1248. 10.1016/j.psyneuen.2011.03.001

28. Hackney, A. C. (2021). Menstrual Cycle Hormonal Changes and Energy Substrate Metabolism in Exercising Women: A Perspective. Int J Environ Res Public Health, 18(19). 10.3390/ijerph181910024

29. Hackney, A. C., Koltun, K. J., & Williett, H. N. (2022). Menstrual cycle hormonal changes: estradiol-β-17 and progesterone interactions on exercise fat oxidation. Endocrine, 76(1), 240–242. 10.1007/s12020-022-02998-w

30. Hackney, A. C., McCracken-Compton, M. A., & Ainsworth, B. (1994). Substrate responses to submaximal exercise in the midfollicular and midluteal phases of the menstrual cycle. Int J Sport Nutr, 4(3), 299–308. 10.1123/ijsn.4.3.299

31. Hardy, C. J., & Rejeski, W. J. (1989). Not what, but how one feels: The measurement of affect during exercise. Journal of Sport & Exercise Psychology, 11(3), 304–317.

32. Hatta, H., Atomi, Y., Shinohara, S., Yamamoto, Y., & Yamada, S. (1988). The effects of ovarian hormones on glucose and fatty acid oxidation during exercise in female ovariectomized rats. Horm Metab Res, 20(10), 609–611. 10.1055/s-2007-1010897

33. Hirnstein, M., Stuebs, J., Moè, A., & Hausmann, M. (2023). Sex/Gender Differences in Verbal Fluency and Verbal-Episodic Memory: A Meta-Analysis. Perspectives on Psychological Science, 18(1), 67–90. 10.1177/17456916221082116

34. Isacco, L., Thivel, D., Pelle, A. M., Zouhal, H., Duclos, M., Duche, P., & Boisseau, N. (2012). Oral contraception and energy intake in women: Impact on substrate oxidation during exercise. Applied Physiology, Nutrition, and Metabolism, 37(4), 646–656. 10.1139/h2012-031 %M 22607658

35. Iversen, L., Fielding, S., Lidegaard, Ø., Mørch, L. S., Skovlund, C. W., & Hannaford, P. C. (2018). Association between contemporary hormonal contraception and ovarian cancer in women of reproductive age in Denmark: prospective, nationwide cohort study. Bmj, 362, k3609. 10.1136/bmj.k3609

36. Daniels, K., Mosher, W.D., Jones, J (2013). *Contraceptive Methods Women Have Ever Used: United States, 1982–*2010. https://www.cdc.gov/nchs/data/nhsr/nhsr062.pdf

37. Jasienski, M., & Bazzaz, F. A. (1999). The fallacy of ratios and the testability of models in biology. Oikos, 84, 321–326.

38. Jeyakumar, M., Carlson, K. E., Gunther, J. R., & Katzenellenbogen, J. A. (2011). Exploration of dimensions of estrogen potency: parsing ligand binding and coactivator binding affinities. J Biol Chem, 286(15), 12971–12982. 10.1074/jbc.M110.205112

39. Jones, A. M. (1998). A five year physiological case study of an Olympic runner. Br J Sports Med, 32(1), 39–43. 10.1136/bjsm.32.1.39

40. Kilpatrick, M., Kraemer, R., Bartholomew, J., Acevedo, E., & Jarreau, D. (2007). Affective responses to exercise are dependent on intensity rather than total work. Med Sci Sports Exerc, 39(8), 1417–1422. 10.1249/mss.0b013e31806ad73c

41. Kirschbaum, C., Platte, P., Pirke, K.-M., & Hellhemmer, D. (1996). Adrenocortical activation following stressful exercise: Further evidence for attenuated free cortisol responses in women using oral contraceptives. Stress Medicine, 12(3), 137–143. 10.1002/(SICI)1099-1700(199607)12:3<137::AID-SMI685>3.0.CO;2-C

42. Kuhnz, W., al-Yacoub, G., & Fuhrmeister, A. (1992). Pharmacokinetics of levonorgestrel and ethinylestradiol in 9 women who received a low-dose oral contraceptive over a treatment period of 3 months and, after a wash-out phase, a single oral administration of the same contraceptive formulation. Contraception, 46(5), 455–469. 10.1016/0010-7824(92)90149-n

43. Lee, V., Rutherfurd-Markwick, K., & Ali, A. (2018). Effect of carbohydrate ingestion during cycling exercise on affective valence and activation in recreational exercisers. Journal of Sports Sciences, 36(3), 340–347.

44. Louw-du Toit, R., Perkins, M. S., Hapgood, J. P., & Africander, D. (2017). Comparing the androgenic and estrogenic properties of progestins used in contraception and hormone therapy. Biochem Biophys Res Commun, 491(1), 140–146. 10.1016/j.bbrc.2017.07.063

45. Luine, V. N. (2014). Estradiol and cognitive function: past, present and future. Horm Behav, 66(4), 602–618. 10.1016/j.yhbeh.2014.08.011

46. Magkos, F., Fabbrini, E., Patterson, B. W., Mittendorfer, B., & Klein, S. (2022). Physiological interindividual variability in endogenous estradiol concentration does not influence adipose tissue and hepatic lipid kinetics in women. Eur J Endocrinol, 187(3), 391–398. 10.1530/eje-22-0410

47. Maki, P. M., Rich, J. B., & Rosenbaum, R. S. (2002). Implicit memory varies across the menstrual cycle: estrogen effects in young women. Neuropsychologia, 40(5), 518–529. 10.1016/s0028-3932(01)00126-9

48. Marcora, S. M., & Staiano, W. (2010). The limit to exercise tolerance in humans: mind over muscle? Eur J Appl Physiol, 109(4), 763–770. 10.1007/s00421-010-1418-6

49. Martin, D., Sale, C., Cooper, S. B., & Elliott-Sale, K. J. (2018). Period Prevalence and Perceived Side Effects of Hormonal Contraceptive Use and the Menstrual Cycle in Elite Athletes. Int J Sports Physiol Perform, 13(7), 926–932. 10.1123/ijspp.2017-0330

50. McNulty, K. L., Elliott-Sale, K. J., Dolan, E., Swinton, P. A., Ansdell, P., Goodall, S., Thomas, K., & Hicks, K. M. (2020). The Effects of Menstrual Cycle Phase on Exercise Performance in Eumenorrheic Women: A Systematic Review and Meta-Analysis. Sports Med, 50(10), 1813–1827. 10.1007/s40279-020-01319-3

51. Merrell, L. H., Perkin, O. J., Bradshaw, L., Collier-Bain, H. D., Collins, A. J., Davies, S., Eddy, R., Hickman, J. A., Nicholas, A. P., Rees, D., Spellanzon, B., James, L. J., McKay, A. K. A., Smith, H. A., Turner, J. E., Koumanov, F., Maher, J., Thompson, D., Gonzalez, J. T., & Betts, J. A. (2024). Myths and Methodologies: Standardisation in human physiology research-should we control the controllables? Exp Physiol, 109(7), 1099–1108. 10.1113/ep091557

52. Mitchell, V. E., & Welling, L. L. M. (2020). Not all progestins are created equally: Considering unique progestins individually in psychobehavioral research. Adaptive Human Behavior and Physiology, 6(3), 381–412. 10.1007/s40750-020-00137-1

53. Mordecai, K. L., Rubin, L. H., & Maki, P. M. (2008). Effects of menstrual cycle phase and oral contraceptive use on verbal memory. Horm Behav, 54(2), 286–293. 10.1016/j.yhbeh.2008.03.006

54. Munive, V., Santi, A., & Torres-Aleman, I. (2016). A Concerted Action Of Estradiol And Insulin Like Growth Factor I Underlies Sex Differences In Mood Regulation By Exercise. Scientific Reports, 6(1), 25969. 10.1038/srep25969

55. Oosthuyse, T., Strauss, J. A., & Hackney, A. C. (2023). Understanding the female athlete: molecular mechanisms underpinning menstrual phase differences in exercise metabolism. European Journal of Applied Physiology, 123(3), 423–450. 10.1007/s00421-022-05090-3

56. Ortega-Santos, C. P., Barba-Moreno, L., Cupeiro, R., & Peinado Lozano, A. B. Substrate oxidation in female adults during endurance exercise throughout menstrual cycle phases: IronFEMME pilot study. 10.14198/jhse.2018.133.07

57. Peragine, D., Simeon-Spezzaferro, C., Brown, A., Gervais, N. J., Hampson, E., & Einstein, G. (2020). Sex difference or hormonal difference in mental rotation? The influence of ovarian milieu. Psychoneuroendocrinology, 115, 104488. 10.1016/j.psyneuen.2019.104488

58. Prado, R. C. R., Silveira, R., Kilpatrick, M. W., Pires, F. O., & Asano, R. Y. (2021). The effect of menstrual cycle and exercise intensity on psychological and physiological responses in healthy eumenorrheic women. Physiology & Behavior, 232, 113290. 10.1016/j.physbeh.2020.113290

59. Rey, A. (1941). L’examen psychologique dans les cas d’encéphalopathie traumatique. (Les problems.). [The psychological examination in cases of traumatic encepholopathy. Problems.]. Archives de Psychologie, 28, 215–285.

60. Rhodes, R. E., & Kates, A. (2015). Can the Affective Response to Exercise Predict Future Motives and Physical Activity Behavior? A Systematic Review of Published Evidence. Ann Behav Med, 49(5), 715–731. 10.1007/s12160-015-9704-5

61. Rosenberg, L., & Park, S. (2002). Verbal and spatial functions across the menstrual cycle in healthy young women. Psychoneuroendocrinology, 27(7), 835–841. 10.1016/S0306-4530(01)00083-X

62. Ruby, B. C., Robergs, R. A., Waters, D. L., Burge, M., Mermier, C., & Stolarczyk, L. (1997). Effects of estradiol on substrate turnover during exercise in amenorrheic females. Med Sci Sports Exerc, 29(9), 1160–1169. 10.1097/00005768-199709000-00007

63. Schulz, K. F., Altman, D. G., & Moher, D. (2010). CONSORT 2010 statement: updated guidelines for reporting parallel group randomised trials. Bmj, 340, c332. 10.1136/bmj.c332

64. Solís-Ortiz, S., & Corsi-Cabrera, M. (2008). Sustained attention is favored by progesterone during early luteal phase and visuo-spatial memory by estrogens during ovulatory phase in young women. Psychoneuroendocrinology, 33(7), 989–998. 10.1016/j.psyneuen.2008.04.003

65. Stanczyk, F. Z., Archer, D. F., & Bhavnani, B. R. (2013). Ethinyl estradiol and 17β-estradiol in combined oral contraceptives: pharmacokinetics, pharmacodynamics and risk assessment. Contraception, 87(6), 706–727. 10.1016/j.contraception.2012.12.011

66. Stroop, J. R. (1935). Studies of interference in serial verbal reactions. Journal of Experimental Psychology, 18(6), 643–662. 10.1037/h0054651

67. Svebak, S., & Murgatroyd, S. (1985). Metamotivational dominance: a multimethod validation of reversal theory constructs. Journal of personality and social psychology, 48(1), 107.

68. Tiller, N. B., Elliott-Sale, K. J., Knechtle, B., Wilson, P. B., Roberts, J. D., & Millet, G. Y. (2021). Do Sex Differences in Physiology Confer a Female Advantage in Ultra-Endurance Sport? Sports Med, 51(5), 895–915. 10.1007/s40279-020-01417-2

69. van Heusden, A. M., & Fauser, B. C. (1999). Activity of the pituitary-ovarian axis in the pill-free interval during use of low-dose combined oral contraceptives. Contraception, 59(4), 237–243. 10.1016/s0010-7824(99)00025-6

70. van Hylckama Vlieg, A., Helmerhorst, F. M., Vandenbroucke, J. P., Doggen, C. J., & Rosendaal, F. R. (2009). The venous thrombotic risk of oral contraceptives, effects of oestrogen dose and progestogen type: results of the MEGA case-control study. Bmj, 339, b2921. 10.1136/bmj.b2921

71. Vandenberg, S. G., & Kuse, A. R. (1978). Mental rotations, a group test of three-dimensional spatial visualization. Percept Mot Skills, 47(2), 599–604. 10.2466/pms.1978.47.2.599

72. Voyer, D., Saint Aubin, J., Altman, K., & Gallant, G. (2021). Sex differences in verbal working memory: A systematic review and meta-analysis. Psychol Bull, 147(4), 352–398. 10.1037/bul0000320

73. Warren, A. M., Gurvich, C., Worsley, R., & Kulkarni, J. (2014). A systematic review of the impact of oral contraceptives on cognition. Contraception, 90(2), 111–116. 10.1016/j.contraception.2014.03.015

74. Wenz, M., Berend, J. Z., Lynch, N. A., Chappell, S., & Hackney, A. C. (1997). Substrate oxidation at rest and during exercise: effects of menstrual cycle phase and diet composition. J Physiol Pharmacol, 48(4), 851–860.

75. Wesnes, K., & Warburton, D. M. (1984). Effects of scopolamine and nicotine on human rapid information processing performance. Psychopharmacology (Berl), 82(3), 147–150. 10.1007/bf00427761

76. Westhoff, C. L., Torgal, A. H., Mayeda, E. R., Pike, M. C., & Stanczyk, F. Z. (2010). Pharmacokinetics of a combined oral contraceptive in obese and normal-weight women. Contraception, 81(6), 474–480. 10.1016/j.contraception.2010.01.016

77. Yue, Y., Zhang, C., Zhang, X., Zhang, S., Liu, Q., Hu, F., Lv, X., Li, H., Yang, J., Wang, X., Chen, L., Yao, Z., Duan, H., & Niu, W. (2020). An AMPK/Axin1-Rac1 signaling pathway mediates contraction-regulated glucose uptake in skeletal muscle cells. American Journal of Physiology-Endocrinology and Metabolism, 318(3), E330–E342. 10.1152/ajpendo.00272.2019

78. Zapata, L. B., Steenland, M. W., Brahmi, D., Marchbanks, P. A., & Curtis, K. M. (2013). Effect of missed combined hormonal contraceptives on contraceptive effectiveness: a systematic review. Contraception, 87(5), 685–700. 10.1016/j.contraception.2012.08.035

79. Zderic, T. W., Coggan, A. R., & Ruby, B. C. (2001). Glucose kinetics and substrate oxidation during exercise in the follicular and luteal phases. J Appl Physiol (1985), *90*(2), 447-453. 10.1152/jappl.2001.90.2.447

80. Zehnder, M., Ith, M., Kreis, R., Saris, W., Boutellier, U., & Boesch, C. (2005). Gender-specific usage of intramyocellular lipids and glycogen during exercise. Med Sci Sports Exerc, 37(9), 1517–1524. 10.1249/01.mss.0000177478.14500.7c

